# An ancestry-matched Mendelian randomisation analysis of kidney function and heart failure subtypes in African ancestry populations

**DOI:** 10.64898/2026.07.15.26358145

**Authors:** Ngone D. Gaye, Abdoulaye Diawara

## Abstract

Chronic kidney disease and heart failure disproportionately burden populations of African ancestry, yet Mendelian randomisation (MR) studies of the causal relationship between kidney function and heart failure subtypes have been conducted exclusively in European ancestry populations. We performed a forward two-sample MR analysis to evaluate the causal effect of genetically predicted estimated glomerular filtration rate (eGFR) on heart failure with preserved ejection fraction (HFpEF) and heart failure with reduced ejection fraction (HFrEF) in individuals of African ancestry. Genetic instruments were selected from an African ancestry eGFR genome-wide association study (N = 67,943) at genome-wide significance, with linkage disequilibrium clumping using an African ancestry reference panel. Heart failure subtype summary statistics were obtained from the Million Veteran Program (HFpEF: 5,379 cases / 113,041 controls; HFrEF: 9,104 cases / 109,632 controls). Six independent SNPs (F-statistics 30.5–107.3; R² = 0.62%) were retained as instruments. The primary inverse-variance weighted analysis provided no evidence of a causal effect of eGFR on HFpEF (OR 0.92, 95% CI 0.80– 1.06, p = 0.248) or HFrEF (OR 0.98, 95% CI 0.78–1.23, p = 0.878). Sensitivity analyses were directionally consistent. There was no evidence of heterogeneity or directional pleiotropy. Minimum detectable effects at 80% power were OR 1.28 for HFpEF and OR 1.22 for HFrEF. These null findings should be interpreted as inconclusive given current power constraints; larger ancestry-matched studies are needed.

## Introduction

Chronic kidney disease and heart failure frequently coexist and interact. Indeed, reduced kidney function is independently associated with a substantially elevated risk of incident heart failure, with hazard ratios exceeding two at eGFR below 45 ml/min/1.73m² after adjustment for traditional cardiovascular risk factors, and associations are stronger for HFpEF than HFrEF [1]. Cardiovascular disease, including heart failure, accounts for 40–50% of all deaths in advanced CKD stages [2]. This bidirectional organ crosstalk, formalised as cardiorenal syndrome, operates through interconnected haemodynamic, neurohormonal, and inflammatory pathways [3–5]. Furthermore, this burden is not evenly distributed: populations of African ancestry experience a disproportionate burden of both conditions, reflecting the combined influence of cardiometabolic risk, infectious and inflammatory exposures, structural determinants of health, and ancestry-specific genetic architecture [6–8]. In sub-Saharan Africa, CKD affects an estimated 13–14% of the general population, and conventional cardiovascular risk factors alone fail to account for a substantial proportion of cases, with infectious exposures, environmental factors, and genetic predisposition contributing independently [6]. APOL1 G1 and G2 risk alleles, which reach their highest frequencies in West Africa, confer substantially elevated risk of focal segmental glomerulosclerosis, hypertension-associated kidney failure, and other nephropathies, with even monoallelic carriage recently shown to increase CKD odds in West African cohorts [7, 9, 10]. Yet APOL1 alone does not fully explain the excess burden, pointing instead to a complex, multifactorial risk architecture extending beyond any single gene.

Heart failure disparities in African ancestry populations are equally pronounced. Incident heart failure before age 50 occurred almost exclusively among Black participants in the CARDIA cohort, with cumulative incidences of 1.1% in Black women and 0.9% in Black men, compared to 0.08% and 0% respectively in their White counterparts [11]. Black adults carry the highest age-adjusted heart failure mortality of any racial group, particularly in the 35–64-year age bracket, and African American women have the highest heart failure prevalence of any intersection of race and sex in the United States [12]. In sub-Saharan Africa, the clinical phenotype is distinct: heart failure predominantly affects younger, working-age adults arises from non-ischaemic aetiologies, including hypertensive heart disease, dilated cardiomyopathy, and rheumatic disease, and carries a one-year mortality of approximately 34% in Africa, far exceeding rates reported in high-income countries [13, 14]. Within this landscape, HFpEF deserves particular attention: Black patients are diagnosed at younger ages and carry a heavier burden of its key risk factors such as hypertension, obesity, and diabetes, yet remain profoundly underrepresented in HFpEF trials and genomic studies [8, 15]. Whether the disproportionate CKD burden in these populations translates into a subtype-specific causal effect on heart failure risk, and whether that effect differs between HFpEF and HFrEF, remains poorly characterised. Mendelian randomisation (MR) uses genetic variants as instruments for modifiable exposures to enable causal inference free from confounding and reverse causation [16]. Several MR studies have examined the causal effect of kidney function or CKD on heart failure subtypes, but these have been conducted almost exclusively in European ancestry populations using instruments derived from European genome-wide association study (GWAS), and have largely yielded null findings [17–19]. The transferability of such instruments to African ancestry populations is substantially reduced by differences in linkage disequilibrium structure and allele frequencies [20]. To our knowledge, no MR study has examined the causal effect of kidney function on heart failure subtypes in African ancestry populations using ancestry-matched genetic instruments. We therefore aimed to evaluate the causal effect of kidney function on HFpEF and HFrEF in individuals of African ancestry using a forward two-sample Mendelian randomisation approach.

## Methods

### 1. Study design

We performed a forward two-sample Mendelian randomisation (MR) analysis to investigate whether genetically predicted kidney function has a causal effect on the risk of heart failure subtypes in individuals of African ancestry. In the two-sample MR framework, genetic associations with the exposure (eGFR) and outcome (HF subtypes) are drawn from separate, non-overlapping genome-wide association study (GWAS) samples. Under the three MR assumptions, namely relevance (instruments are robustly associated with the exposure), independence (instruments are not associated with confounders), and exclusion restriction (instruments affect the outcome only through the exposure), this design provides estimates that can be interpreted as causal effects (Figure 1). Analyses were restricted to African-ancestry GWAS throughout, including for linkage disequilibrium (LD) reference panels used during instrument clumping, to preserve ancestry-matched inference and avoid bias from population stratification.

**Figure 1.**
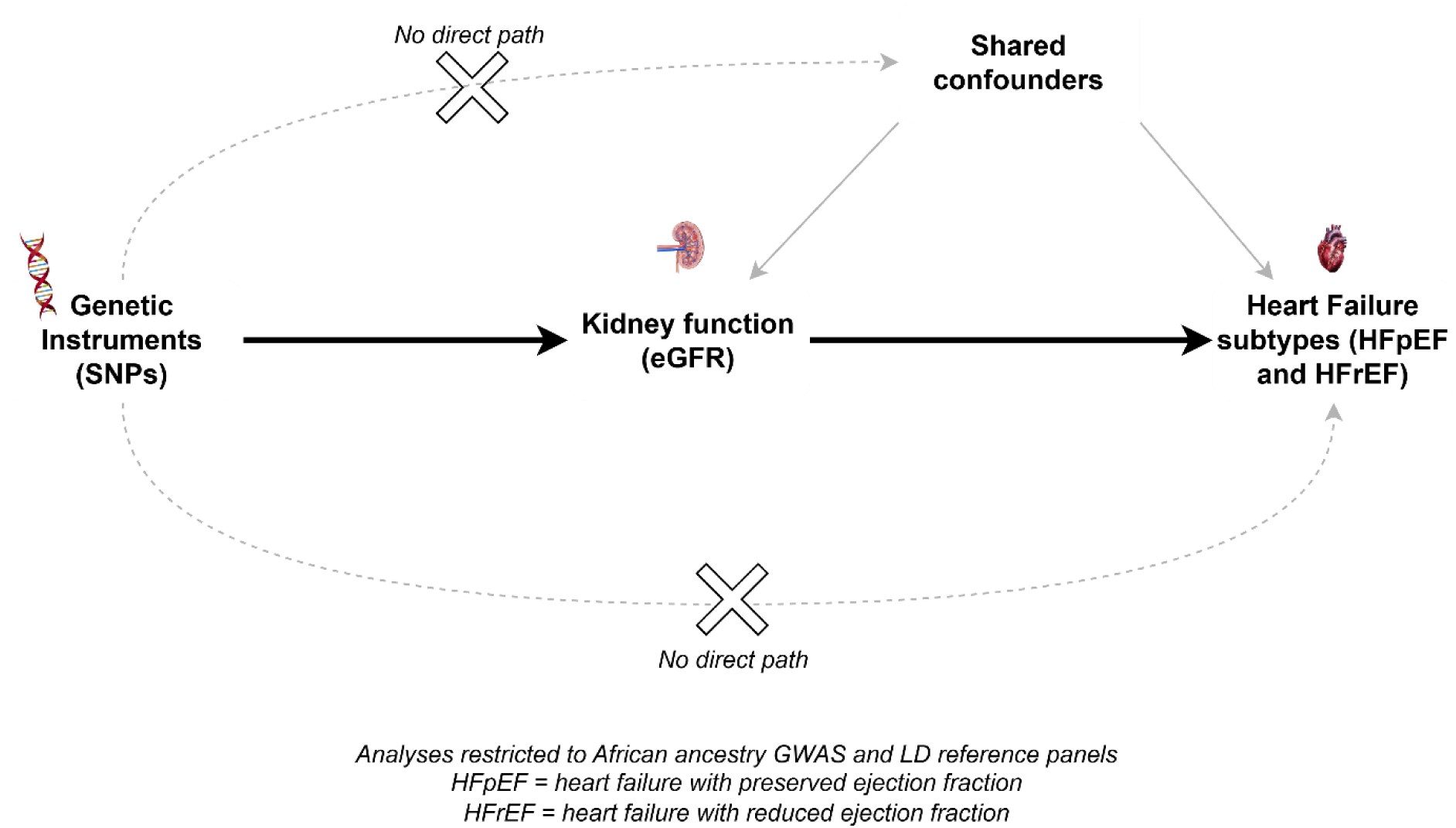
Directed acyclic graph of the two-sample Mendelian randomisation framework Genetic instruments (SNPs) associated with kidney function (eGFR) are used to estimate its causal effect on heart failure subtypes (HFpEF and HFrEF). Valid instruments influence the outcome only through the exposure; the crossed dashed paths indicate the assumed absence of a direct instrument–outcome effect and of instrument–confounder associations. Analyses were restricted to African ancestry GWAS and linkage disequilibrium reference panels throughout. HFpEF, heart failure with preserved ejection fraction; HFrEF, heart failure with reduced ejection fraction.

### 2. Data sources

#### Exposure — kidney function

Genetic associations with estimated glomerular filtration rate based on serum creatinine (eGFRcrea) were obtained from the African ancestry subset of the Kidney Multiome-based Genetic Scorecard GWAS (N = 67,943) [21]. Summary statistics included single-nucleotide polymorphisms (SNPs) identifiers, chromosomal position, effect and other alleles, effect allele frequency (EAF), effect estimates (β), standard errors (SE), and p-values.

#### Outcomes — heart failure subtypes

Heart failure subtype summary statistics were obtained from the Million Veteran Program (MVP) GWAS of African American and Afro-Caribbean participants [22]. Two phenotypes were analysed: heart failure with preserved ejection fraction (HFpEF; PheCode 428.4; GWAS Catalog accession GCST90477963; N_cases_ = 5,379; N_controls_ = 113,041; Neff = 20,539) and heart failure with reduced ejection fraction (HFrEF; PheCode 428.3; GWAS Catalog accession GCST90477961; N_cases_ = 9,104; N_controls_ = 109,632; Neff = 33,624). Effective sample sizes were computed using the harmonic mean formula: N_eff_ = 4 / (1/N_cases_ + 1/N_controls_). As standard errors were not directly available in the deposited MVP summary statistics, they were derived from the provided 95% confidence intervals as SE = (log (CI_upper_) − log (CI_lower_)) / (2 × 1.96) assuming the reported confidence intervals were based on a normal approximation, as is standard practice in large-scale GWAS.

### 3. Instrument selection and strength

Genetic instruments for eGFRcrea were selected at genome-wide significance (p < 5×10^−8^). Summary statistics were first restricted to biallelic SNPs with unambiguous ACGT alleles. Independent instruments were then identified using LD clumping with an African ancestry reference panel (r^2^ < 0.001; 10 Mb window) via the IEU OpenGWAS platform [23]. This stringent r² threshold minimises correlation between instruments, prioritising independence over instrument density given the modest African ancestry GWAS sample size. Instrument strength was assessed using the F-statistic for each SNP, calculated as (β/SE)^2^. SNPs with F < 10 were excluded to minimise weak-instrument bias [24]. The proportion of variance in eGFRcrea explained by retained instruments (R^2^) was computed to contextualise instrument informativeness.

### 4. Harmonisation

Exposure and outcome summary statistics were harmonised using the TwoSampleMR package (v0.6.30) [23] with action = 2, which attempts to align effect alleles across datasets and resolves palindromic SNPs using allele frequency information where available; SNPs with irresolvable strand ambiguity were excluded. An attrition table was recorded at each step from instrument selection through harmonisation to document SNP retention.

### 5. MR analysis

#### Primary analysis

The primary causal effect was estimated using the inverse-variance weighted (IVW) random-effects model, which provides an efficient pooled estimate under the assumption that any horizontal pleiotropy is balanced (InSIDE assumption) [25]. Results are expressed as odds ratios (OR) with 95% confidence intervals per standard deviation (SD) increase in genetically predicted eGFRcrea.

#### Sensitivity analyses

Three complementary estimators were applied to assess robustness to violations of the exclusion restriction assumption. The weighted median estimator yields consistent estimates when at least 50% of instrument weight derives from valid variants [26]. The weighted mode estimator is consistent when the largest subset of instruments share a common causal effect [27]. MR-Egger regression relaxes the InSIDE assumption by allowing an unconstrained intercept; the intercept itself provides a formal test for directional horizontal pleiotropy [28], though its power is limited with small instrument sets and results should be interpreted accordingly. Between-instrument heterogeneity was assessed using Cochran’s Q statistic. Leave-one-out and single-SNP analyses were conducted to detect influential variants. MR-PRESSO was not applied, as outlier detection with this method has low power with fewer than ten instruments [29].

#### Power and minimum detectable effect

To contextualise the precision of estimates, we calculated the minimum detectable effect (MDE) at 80% power and α = 0.05 using the total variance in eGFRcrea explained by retained instruments (R^2^) and the effective outcome sample size, following the approach of Brion et al. [30].

### 6. Software and reproducibility

All analyses were conducted in R version 4.5.0 (R Foundation for Statistical Computing). Key packages used were TwoSampleMR (v0.6.30), ieugwasr, data. table, ggplot2, flextable, and officer. LD clumping was performed using African ancestry reference panels accessed via the IEU OpenGWAS platform.

## Results

### Instrument selection and strength

After applying a genome-wide significance threshold (p < 5×10^−8^), LD clumping using an African ancestry reference panel (r^2^ < 0.001; 10 Mb window), and harmonisation with outcome summary statistics, six independent SNPs were retained as genetic instruments for eGFRcrea in both analyses. SNP attrition at each step is presented in Table 1.

**Table 1.** SNP attrition from instrument selection to analysis.

| Step | HFpEF (n SNPs) | HFrEF (n SNPs) |
| --- | --- | --- |
| GWS instruments (pre-clump) | 195 | 195 |
| Post-clump | 6 | 6 |
| Present in outcome | 6 | 6 |
| Harmonised | 6 | 6 |
| Strong ($F \geq 10$) | 6 | 6 |

*GWS, genome-wide significant ( $p < 5 \times 10^{-8}$ ); AFR, African ancestry linkage disequilibrium reference panel. HFpEF, heart failure with preserved ejection fraction; HFrEF, heart failure with reduced ejection fraction.*

All six instruments were present in both MVP outcome GWAS files with no attrition at harmonisation, reflecting the broad genomic coverage of the MVP platform. All instruments demonstrated adequate strength, with F-statistics ranging from 30.5 to 107.3 (all > 10), indicating low risk of weak-instrument bias (Table 2).

**Table 2.**
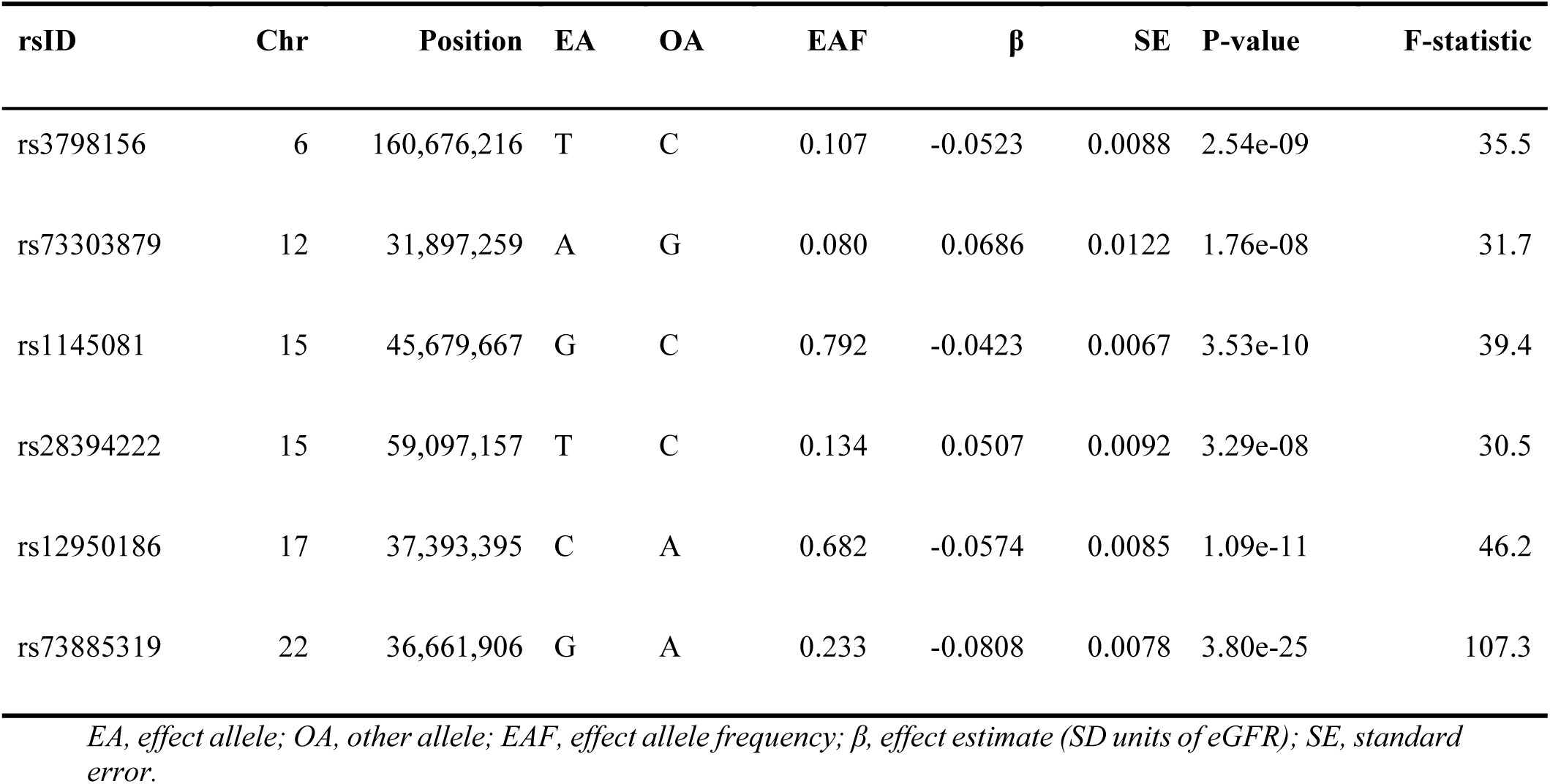
Genetic instruments for estimated glomerular filtration rate (eGFRcrea) in African ancestry populations.

| rsID | Chr | Position | EA | OA | EAF | $\beta$ | SE | P-value | F-statistic |
| --- | --- | --- | --- | --- | --- | --- | --- | --- | --- |
| rs3798156 | 6 | 160,676,216 | T | C | 0.107 | -0.0523 | 0.0088 | 2.54e-09 | 35.5 |
| rs73303879 | 12 | 31,897,259 | A | G | 0.080 | 0.0686 | 0.0122 | 1.76e-08 | 31.7 |
| rs1145081 | 15 | 45,679,667 | G | C | 0.792 | -0.0423 | 0.0067 | 3.53e-10 | 39.4 |
| rs28394222 | 15 | 59,097,157 | T | C | 0.134 | 0.0507 | 0.0092 | 3.29e-08 | 30.5 |
| rs12950186 | 17 | 37,393,395 | C | A | 0.682 | -0.0574 | 0.0085 | 1.09e-11 | 46.2 |
| rs73885319 | 22 | 36,661,906 | G | A | 0.233 | -0.0808 | 0.0078 | 3.80e-25 | 107.3 |

*EA, effect allele; OA, other allele; EAF, effect allele frequency; $\beta$ , effect estimate (SD units of eGFR); SE, standard error.*

The retained instruments collectively explained 0.62% of the variance in eGFRcrea (R^2^ = 0.00617). Steiger filtering confirmed that all six SNPs explained greater variance in eGFRcrea than in either outcome (all p < 0.01), supporting the hypothesised causal direction and retaining all instruments for analysis (Additional file 1: Table S1).

### eGFR → HFpEF

The primary IVW random-effects analysis provided no evidence of a causal effect of genetically predicted eGFRcrea on HFpEF risk (OR 0.92, 95% CI 0.80–1.06, p = 0.248; 6 SNPs) (Figure 2). Sensitivity analyses were directionally consistent with the primary estimate across all three estimators: weighted median (OR 0.88, 95% CI 0.56–1.37, p = 0.570), MR-Egger (OR 0.50, 95% CI 0.09–2.61, p = 0.455), and weighted mode (OR 0.85, 95% CI 0.36–2.00, p = 0.720). Full numerical results for all MR methods are provided in Additional file 1: Table S2. Scatter plots showed a consistent negative slope across estimators, directionally consistent with higher genetically predicted eGFR being associated with lower HFpEF risk, though confidence intervals were wide and estimates did not reach statistical significance (Additional file 2: Figure S1a). There was no evidence of between-instrument heterogeneity (Cochran’s Q p = 0.983) or directional horizontal pleiotropy (MR-Egger intercept 0.038, SE 0.051, p = 0.496), though the power of the Egger intercept test is limited with six instruments and this result should be interpreted cautiously (Additional file 1: Table S3). Leave-one-out analysis confirmed that no single instrument drove the overall estimate (Additional file 2: Figure S2a).

**Figure 2.**
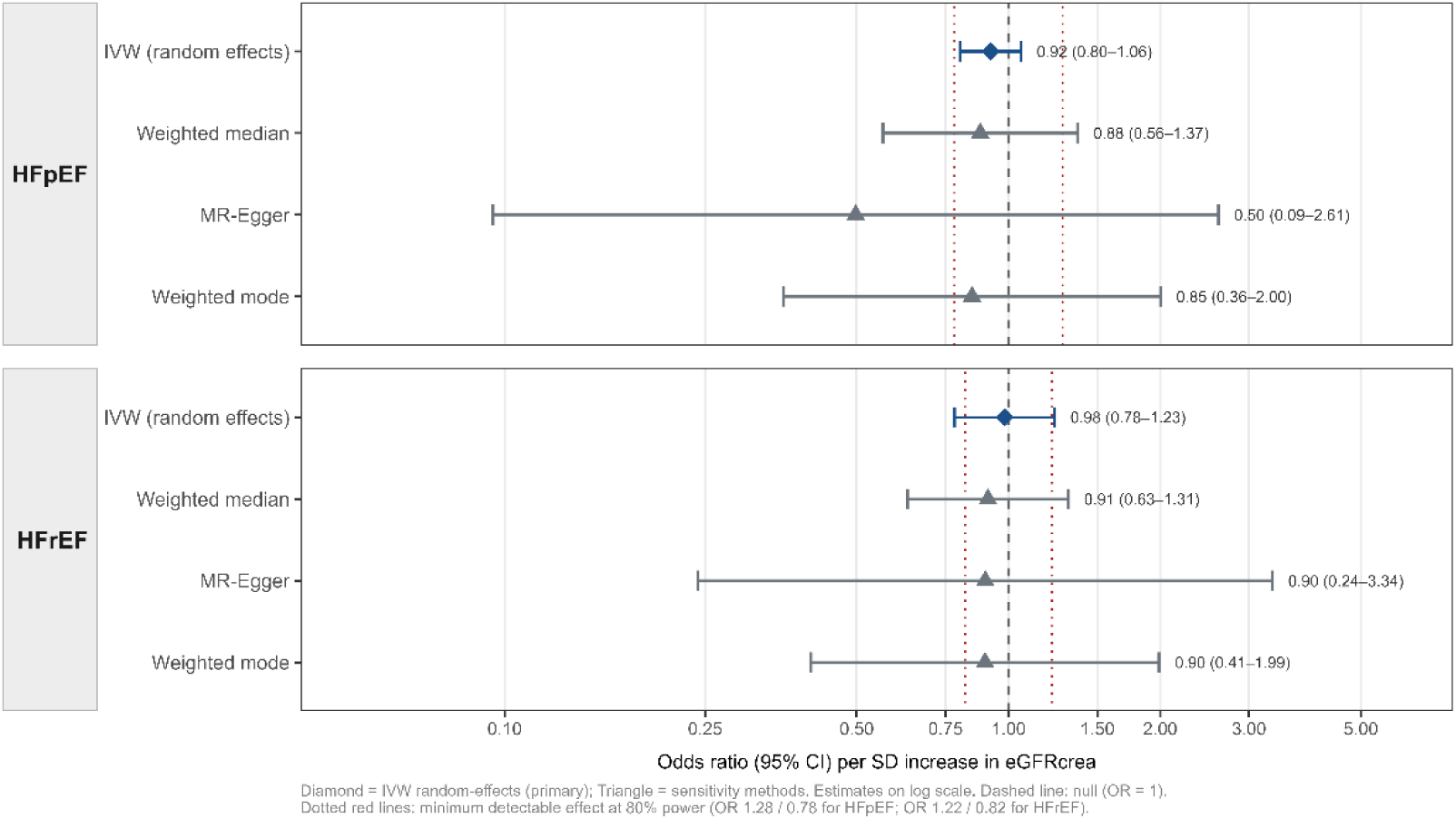
Mendelian randomisation estimates for genetically predicted eGFRcrea on heart failure subtypes. Results are shown for HFpEF (upper panel) and HFrEF (lower panel) in African ancestry populations. Odds ratios are per standard deviation increase in genetically predicted eGFRcrea. CI, confidence interval; HFpEF, heart failure with preserved ejection fraction; HFrEF, heart failure with reduced ejection fraction; IVW, inverse-variance weighted; MDE, minimum detectable effect; MR, Mendelian randomisation.

### eGFR → HFrEF

The primary IVW random-effects analysis similarly showed no evidence of a causal effect of genetically predicted eGFRcrea on HFrEF risk (OR 0.98, 95% CI 0.78–1.23, p = 0.878; 6 SNPs) (Figure 2). Results were consistent across sensitivity estimators: weighted median (OR 0.91, 95% CI 0.63–1.31, p = 0.616), MR-Egger (OR 0.90, 95% CI 0.24–3.34, p = 0.882), and weighted mode (OR 0.90, 95% CI 0.41–1.99, p = 0.802). In contrast to HFpEF, scatter plots showed a near-horizontal slope across estimators, providing no directional signal in either direction (Additional file 2: Figure S1b). There was no evidence of between-instrument heterogeneity (Cochran’s Q p = 0.693); the MR-Egger intercept test did not detect statistically significant directional pleiotropy (intercept 0.005, SE 0.040, p = 0.899), though power is limited with six instruments (Additional file 1: Table S3). Leave-one-out analysis confirmed robustness of the null estimate to individual instrument removal (Additional file 2: Figure S2b). Funnel plots for both outcomes are provided in Additional file 2 (Figure S3).

### Power and minimum detectable effects

At 80% power and α = 0.05, the minimum detectable effect corresponded to OR 1.28 for HFpEF and OR 1.22 for HFrEF (Additional file 1: Table S4).

## Discussion

In this two-sample Mendelian randomisation study using African ancestry-specific genome-wide association data, we found no evidence supporting a statistically significant causal effect of genetically predicted kidney function on HFpEF or HFrEF risk. Across MR estimators, estimates for HFpEF were consistently below the null, compatible with a directionally protective association of genetically predicted higher eGFR with HFpEF risk, although confidence intervals crossed the null value (IVW OR 0.92, 95% CI 0.80–1.06). By contrast, estimates for HFrEF were close to unity (IVW OR 0.98, 95% CI 0.78–1.23). There was no evidence of heterogeneity or directional pleiotropy. These findings should be interpreted considering limited statistical power, as the minimum detectable effects at 80% power corresponded to ORs of 1.28 for HFpEF and 1.22 for HFrEF. Nevertheless, our study extends the cardiorenal MR literature to African ancestry populations, where genetic epidemiological evidence remains limited despite the substantial burden of both chronic kidney disease and heart failure.

Prior MR studies of kidney function and heart failure have been conducted predominantly in European ancestry populations and have yielded largely null findings for the forward direction [17, 31]. Only one of these studies examined HFpEF and HFrEF separately, reporting null findings for both subtypes [17]. Together with our findings, this pattern suggests that genetically proxied kidney function may not exert a large independent causal effect on heart failure risk, although smaller effects cannot be excluded. The observed epidemiological association between CKD and heart failure may therefore be driven, at least in part, by shared cardiometabolic risk factors, haemodynamic stress, inflammation, treatment effects, and reverse causation. Whether these pathways operate similarly across ancestry groups remains uncertain, particularly in African ancestry populations, where CKD aetiology, genetic architecture, and heart failure phenotypes differ markedly.

The directional contrast observed between HFpEF and HFrEF in our study warrants cautious biological interpretation. Although neither subtype-specific estimate reached conventional statistical significance, the consistently negative direction of effect across MR estimators for HFpEF, contrasted with the near-null pattern for HFrEF is hypothesis-generating and warrants brief biological contextualisation. HFpEF is increasingly conceptualised as a systemic, comorbidity-driven syndrome in which inflammation, endothelial dysfunction, microvascular impairment [32], and cardiorenal interactions contribute to myocardial stiffness and diastolic dysfunction [33, 34]. In this context, impaired kidney function may promote HFpEF-relevant pathways through sodium and volume retention, neurohormonal activation, sympathetic overactivity, endothelial dysfunction, and left ventricular hypertrophy [35–37]. These mechanisms provide a plausible framework for interpreting why genetically predicted higher eGFR showed a more consistently inverse direction of association with HFpEF than with HFrEF in our analyses. By contrast, HFrEF is more commonly driven by ischaemic myocardial injury, cardiomyocyte loss, adverse remodelling, and replacement fibrosis, pathways that may be less directly captured by genetically proxied variation in kidney function [37, 38]. Taken together, our findings are consistent with the hypothesis that kidney function may contribute more strongly to the HFpEF biological milieu than to HFrEF. However, given the wide confidence intervals and limited subtype-specific power, this interpretation should be regarded as exploratory and requires confirmation in larger ancestry-matched datasets with sufficient power to formally test heterogeneity by heart failure subtype.

This study is, to our knowledge, the first Mendelian randomisation investigation of kidney function on heart failure subtypes conducted exclusively using African ancestry GWAS data. By restricting both exposure and outcome GWAS to individuals of African ancestry and using ancestry-matched LD reference panels throughout, we avoid the cross-ancestry instrument mismatch that commonly affects MR analyses in non-European populations. The consistency of results across weighted median, MR-Egger, and weighted mode estimators lends further internal coherence to the null findings. These strengths notwithstanding, several limitations merit consideration. The principal limitation of this study is the small instrument set: six SNPs collectively explaining 0.6% of eGFR variance. This constrains the analysis in two related ways. First, statistical power is limited: at 80% power and α = 0.05, minimum detectable effects were OR 1.28 for HFpEF and OR 1.22 for HFrEF; any true causal effect below these thresholds would not have been reliably detected. Second, MR-Egger and weighted median estimators may underperform with small, low-R² instrument sets even when individual F-statistics exceed 10. Additionally, while the exposure and outcome GWAS derive from different consortia, partial sample overlap cannot be fully excluded, although the IVW random-effects method is robust to modest overlap when standard errors are appropriately estimated. The null results reported here should therefore be interpreted as inconclusive rather than as evidence of absence. Resolving this uncertainty will require subtype-stratified MR in adequately powered cohorts, incorporating additional renal traits such as UACR and cystatin-C based eGFR, and exploring effect modification by APOL1 risk genotypes. Such advances will be greatly facilitated by continued investment in continental African genomic infrastructure and cross-consortium collaboration, as exemplified by initiatives such as KidneyGenAfrica [39], whose recent pan-African eGFR meta-analysis represents an important step toward the sample sizes needed to power future MR investigations in this population.

## Conclusion

This study provides the first Mendelian randomisation evidence on the causal effect of kidney function on heart failure subtypes derived exclusively from African ancestry genome-wide association data. We found no statistically significant causal effect of genetically predicted eGFR on either HFpEF or HFrEF risk, a pattern consistent with prior European ancestry MR studies. Given the limited statistical power of the current analysis, these null findings should be interpreted as inconclusive rather than as evidence of absence. Adequately powered, ancestry-matched studies incorporating a broader repertoire of renal traits and larger African ancestry cohorts will be necessary to resolve whether kidney function exerts a subtype-specific causal effect on heart failure risk in this population.

## Abbreviations

CI: confidence interval
CKD: chronic kidney disease
EA: effect allele
EAF: effect allele frequency
eGFR: estimated glomerular filtration rate
eGFRcrea: estimated glomerular filtration rate based on serum creatinine
GWAS: genome-wide association study
GWS: genome-wide significance
HF: heart failure
HFpEF: heart failure with preserved ejection fraction
HFrEF: heart failure with reduced ejection fraction
IEU: Integrative Epidemiology Unit
InSIDE: INstrument Strength Independent of Direct Effect
IVW: inverse-variance weighted
LD: linkage disequilibrium
MDE: minimum detectable effect
MR: Mendelian randomisation
MR: Egger Mendelian randomisation-Egger
MVP: Million Veteran Program
OA: other allele
OR: odds ratio
SD: standard deviation
SE: standard error
SNPs: single-nucleotide polymorphisms
UACR: Urine Albumin-Creatinine Ratio

## Ethics approval and consent to participate

Not applicable. This study used only publicly available genome-wide association study summary statistics. No individual-level human data were accessed, and no ethical approval was required.

## Consent for publication

Not applicable.

## Data availability

The exposure GWAS summary statistics (eGFR, African ancestry, N = 67,943) are publicly available via figshare at https://figshare.com/ndownloader/files/47668984. The outcome GWAS summary statistics are available from the GWAS Catalog under accession numbers GCST90477963 (HFpEF) and GCST90477961 (HFrEF) and are publicly accessible without restrictions. Full analysis code, including configuration files, functions, and analysis scripts, is publicly available at https://github.com/NgoneGaye/HF-MVP-MR to ensure complete reproducibility.

## Authors’ contributions

**Conceptualization:** N.D.G. and A.D.

**Formal analysis:** N.D.G.

**Methodology:** N.D.G.

**Visualization:** N.D.G.

**Writing - original draft:** N.D.G.

**Writing - review & editing:** N.D.G. and A.D.

## Acknowledgements

The authors thank the participants and investigators of the contributing GWAS studies, including the Kidney Multiome-based Genetic Scorecard consortium and the Million Veteran Program, for making summary statistics publicly available.

## References

1. Cheng RK, Roberts MB, Bansal N, Reding K, Salahuddin T, Mamas M, et al. Association of Kidney Function with Incident Heart Failure: An Analysis of the Women’s Health Initiative. J Am Heart Assoc. 2025;14: e037051. 10.1161/JAHA.124.037051.

2. Jankowski J, Floege J, Fliser D, Böhm M, Marx N. Cardiovascular Disease in Chronic Kidney Disease: Pathophysiological Insights and Therapeutic Options. Circulation. 2021 ;143 :1157–72. 10.1161/CIRCULATIONAHA.120.050686.

3. Rangaswami J, Bhalla V, Blair JEA, Chang TI, Costa S, Lentine KL, et al. Cardiorenal Syndrome: Classification, Pathophysiology, Diagnosis, and Treatment Strategies: A Scientific Statement from the American Heart Association. Circulation. 2019;139: e840–78. 10.1161/CIR.0000000000000664.

4. Ronco C, McCullough P, Anker SD, Anand I, Aspromonte N, Bagshaw SM, et al. Cardio-renal syndromes: report from the consensus conference of the acute dialysis quality initiative. Eur Heart J. 2010; 31:703–11. 10.1093/eurheartj/ehp507.

5. Ronco C, Haapio M, House AA, Anavekar N, Bellomo R. Cardiorenal syndrome. J Am Coll Cardiol. 2008; 52:1527–39. 10.1016/j.jacc.2008.07.051.

6. Stanifer JW, Jing B, Tolan S, Helmke N, Mukerjee R, Naicker S, et al. The epidemiology of chronic kidney disease in sub-Saharan Africa: a systematic review and meta-analysis. The Lancet Global Health. 2014;2: e174–81. 10.1016/S2214-109X(14)70002-6.

7. Genovese G, Friedman DJ, Ross MD, Lecordier L, Uzureau P, Freedman BI, et al. Association of trypanolytic ApoL1 variants with kidney disease in African Americans. Science. 2010; 329:841–5. 10.1126/science.1193032.

8. Sirugo G, Williams SM, Tishkoff SA. The Missing Diversity in Human Genetic Studies. Cell. 2019; 177:26–31. 10.1016/j.cell.2019.02.048.

9. Limou S, Nelson GW, Kopp JB, Winkler CA. APOL1 Kidney Risk Alleles: Population Genetics and Disease Associations. Advances in Chronic Kidney Disease. 2014; 21:426–33. 10.1053/j.ackd.2014.06.005.

10. Gbadegesin RA, Ulasi I, Ajayi S, Raji Y, Olanrewaju T, Osafo C, et al. APOL1 Bi- and Monoallelic Variants and Chronic Kidney Disease in West Africans. N Engl J Med. 2025; 392:228–38. 10.1056/NEJMoa2404211.

11. Bibbins-Domingo K, Pletcher MJ, Lin F, Vittinghoff E, Gardin JM, Arynchyn A, et al. Racial Differences in Incident Heart Failure among Young Adults. N Engl J Med. 2009; 360:1179–90. 10.1056/NEJMoa0807265.

12. Piña IL, Jimenez S, Lewis EF, Morris AA, Onwuanyi A, Tam E, et al. Race and Ethnicity in Heart Failure. Journal of the American College of Cardiology. 2021; 78:2589–98. 10.1016/j.jacc.2021.06.058.

13. Damasceno A, Mayosi BM, Sani M, Ogah OS, Mondo C, Ojji D, et al. The Causes, Treatment, and Outcome of Acute Heart Failure in 1006 Africans from 9 Countries: Results of the Sub-Saharan Africa Survey of Heart Failure. Arch Intern Med. 2012; 172:1386. 10.1001/archinternmed.2012.3310.

14. Dokainish H, Teo K, Zhu J, Roy A, AlHabib KF, ElSayed A, et al. Global mortality variations in patients with heart failure: results from the International Congestive Heart Failure (INTER-CHF) prospective cohort study. The Lancet Global Health. 2017;5: e665–72. 10.1016/S2214-109X(17)30196-1.

15. Brown S, Biswas D, Wu J, Ryan M, Bernstein BS, Fairhurst N, et al. Race- and Ethnicity-Related Differences in Heart Failure with Preserved Ejection Fraction Using Natural Language Processing. JACC: Advances. 2024; 3:101064. 10.1016/j.jacadv.2024.101064.

16. Davey Smith G, Hemani G. Mendelian randomization: genetic anchors for causal inference in epidemiological studies. Human Molecular Genetics. 2014;23: R89–98. 10.1093/hmg/ddu328.

17. Lin M, Guo J, Tao H, Gu Z, Tang W, Zhou F, et al. Circulating mediators linking cardiometabolic diseases to HFpEF: a mediation Mendelian randomization analysis. Cardiovasc Diabetol. 2025; 24:201. 10.1186/s12933-025-02738-0.

18. Zhang J, Hu Z, Tan Y, Ye J. Causal relationship from heart failure to kidney function and CKD: A bidirectional two-sample mendelian randomization study. PLoS ONE. 2023;18:e0295532. 10.1371/journal.pone.0295532.

19. Zhou X, Ruan W, Zhao L, Lin K, Li J, Liu H, et al. Causal Links Between Renal Function and Cardiac Structure, Function, and Disease Risk. Glob Heart. 2024; 19:83. 10.5334/gh.1366.

20. Hou L, Wu S, Yuan Z, Xue F, Li H. TEMR: Trans-ethnic mendelian randomization method using large-scale GWAS summary datasets. The American Journal of Human Genetics. 2025; 112:28–43. 10.1016/j.ajhg.2024.11.006.

21. Liu H, Abedini A, Ha E, Ma Z, Sheng X, Dumoulin B, et al. Kidney multiome-based genetic scorecard reveals convergent coding and regulatory variants. Science. 2025 ;387 : eadp4753. 10.1126/science.adp4753.

22. Verma A, Huffman JE, Rodriguez A, Conery M, Liu M, Ho Y-L, et al. Diversity and scale: Genetic architecture of 2068 traits in the VA Million Veteran Program. Science. 2024;385: eadj1182. 10.1126/science.adj1182.

23. Hemani G, Zheng J, Elsworth B, Wade KH, Haberland V, Baird D, et al. The MR-Base platform supports systematic causal inference across the human phenome. eLife. 2018;7: e34408. 10.7554/eLife.34408.

24. Staiger D, Stock JH. Instrumental Variables Regression with Weak Instruments. Econometrica. 1997; 65:557. 10.2307/2171753.

25. Burgess S, Butterworth A, Thompson SG. Mendelian Randomization Analysis with Multiple Genetic Variants Using Summarized Data. Genetic Epidemiology. 2013; 37:658–65. 10.1002/gepi.21758.

26. Bowden J, Davey Smith G, Haycock PC, Burgess S. Consistent Estimation in Mendelian Randomization with Some Invalid Instruments Using a Weighted Median Estimator. Genetic Epidemiology. 2016; 40:304–14. 10.1002/gepi.21965.

27. Hartwig FP, Davey Smith G, Bowden J. Robust inference in summary data Mendelian randomization via the zero modal pleiotropy assumption. International Journal of Epidemiology. 2017; 46:1985–98. 10.1093/ije/dyx102.

28. Bowden J, Davey Smith G, Burgess S. Mendelian randomization with invalid instruments: effect estimation and bias detection through Egger regression. International Journal of Epidemiology. 2015; 44:512–25. 10.1093/ije/dyv080.

29. Verbanck M, Chen C-Y, Neale B, Do R. Detection of widespread horizontal pleiotropy in causal relationships inferred from Mendelian randomization between complex traits and diseases. Nat Genet. 2018; 50:693–8. 10.1038/s41588-018-0099-7.

30. Brion M-JA, Shakhbazov K, Visscher PM. Calculating statistical power in Mendelian randomization studies. International Journal of Epidemiology. 2013; 42:1497–501. 10.1093/ije/dyt179.

31. Hu C, Li Y, Qian Y, Wu Z, Hu B, Peng Z. Kidney function and cardiovascular diseases: a large-scale observational and Mendelian randomization study. Front Immunol. 2023; 14:1190938. 10.3389/fimmu.2023.1190938.

32. Paulus WJ, Tschöpe C. A Novel Paradigm for Heart Failure with Preserved Ejection Fraction: Comorbidities Drive Myocardial Dysfunction and Remodeling Through Coronary Microvascular Endothelial Inflammation. Journal of the American College of Cardiology. 2013; 62:263–71. 10.1016/j.jacc.2013.02.092.

33. ter Maaten JM, Damman K, Verhaar MC, Paulus WJ, Duncker DJ, Cheng C, et al. Connecting heart failure with preserved ejection fraction and renal dysfunction: the role of endothelial dysfunction and inflammation. European Journal of Heart Failure. 2016; 18:588– 98. 10.1002/ejhf.497.

34. Patel RN, Sharma A, Prasad A, Bansal S. Heart Failure with Preserved Ejection Fraction With CKD: A Narrative Review of a Multispecialty Disorder. Kidney Medicine. 2023;5. 10.1016/j.xkme.2023.100705.

35. Akwo EA, Robinson-Cohen C. Mendelian randomization and the association of fibroblast growth factor-23 with heart failure with preserved ejection fraction. Curr Opin Nephrol Hypertens. 2023; 32:305–12. 10.1097/MNH.0000000000000888.

36. van de Wouw J, Broekhuizen M, Sorop O, Joles JA, Verhaar MC, Duncker DJ, et al. chronic kidney disease as a Risk Factor for Heart Failure with Preserved Ejection Fraction: A Focus on Microcirculatory Factors and Therapeutic Targets. Front Physiol. 2019; 10:1108. 10.3389/fphys.2019.01108.

37. Noels H, van der Vorst EPC, Rubin S, Emmett A, Marx N, Tomaszewski M, et al. Renal-Cardiac Crosstalk in the Pathogenesis and Progression of Heart Failure. Circulation Research. 2025; 136:1306–34. 10.1161/CIRCRESAHA.124.325488.

38. Kozman K, Ferrannini G, Benson L, Dahlström U, Hage C, Savarese G, et al. Etiology of Heart Failure Across the Ejection Fraction Spectrum and Association with Prognosis. JACC: Heart Failure. 2025; 13:102491. 10.1016/j.jchf.2025.03.037.

39. Kamiza AB, Chikowore T, Chen G, Ojewunmi O, Machipisa T, Zhou F, et al. KidneyGenAfrica multi-cohort Genome-wide association study and polygenic prediction of kidney function in 110,000 Africans. Nat Commun. 2026; 17:2599. 10.1038/s41467-026-69367-3.

