## Supplemental Figures for "An ancestry-matched Mendelian randomisation analysis of kidney function and heart failure subtypes in African ancestry populations"

**Figure S1. Scatter plots of SNP effects on eGFR and heart failure subtypes**

**Figure S1a. eGFR –> HFpEF**
Scatter plot

**Figure S1b. eGFR –> HFrEF**
Scatter plot

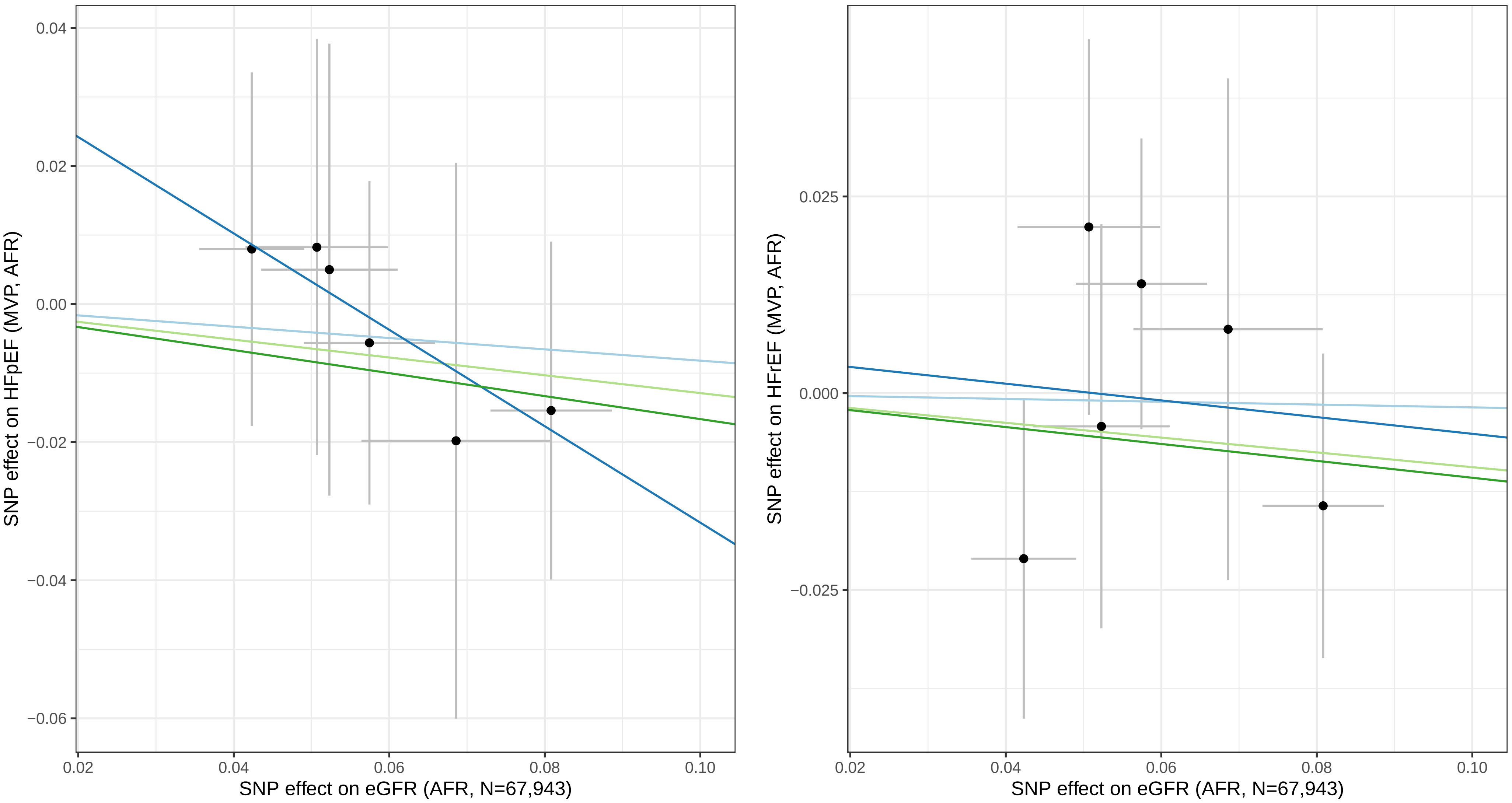


Each point represents a single SNP instrument. Lines show MR method estimates.
IVW = inverse−variance weighted (primary); sensitivity methods shown for comparison.
HFpEF, heart failure with preserved ejection fraction; HFrEF, heart failure with reduced ejection fraction.

**Figure S2. Leave−one−out sensitivity analyses**

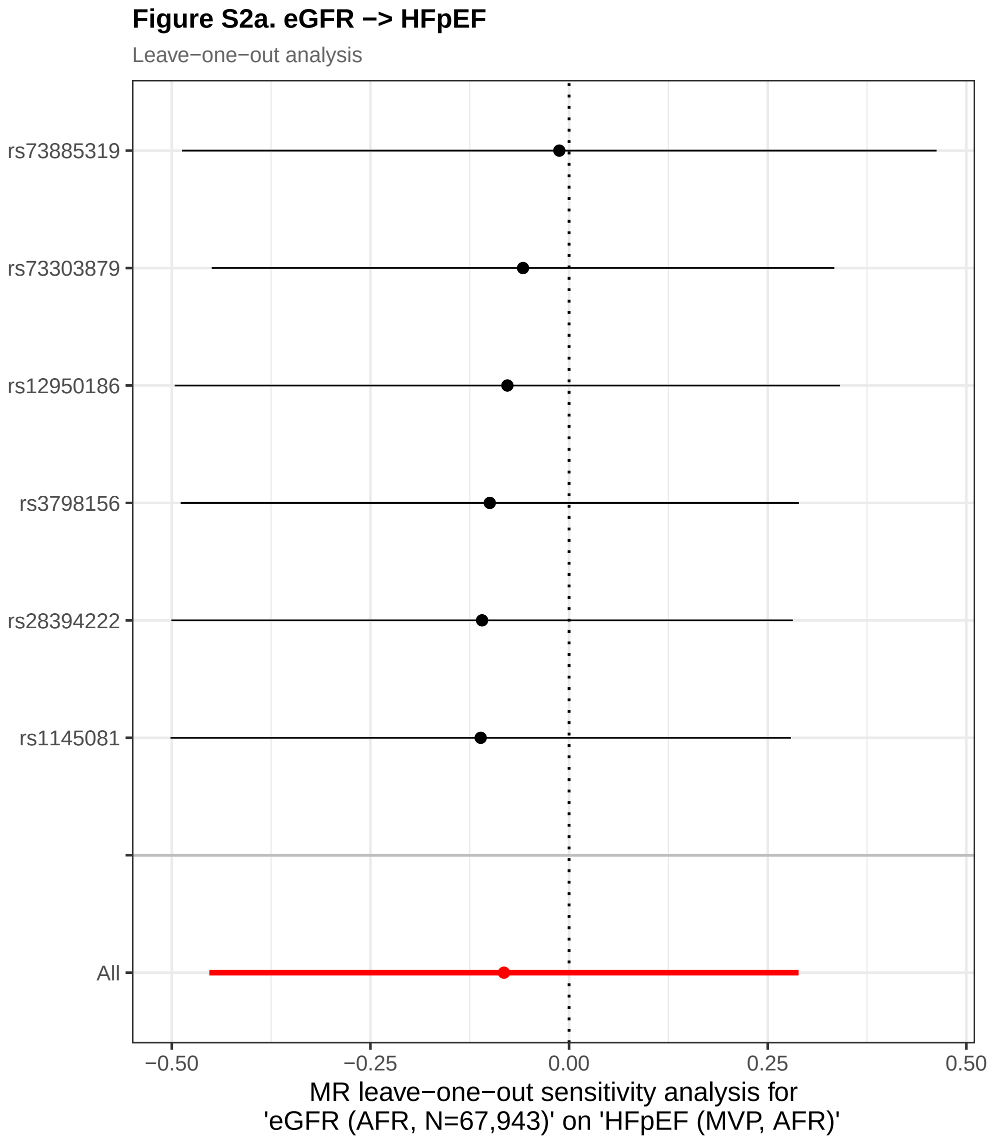


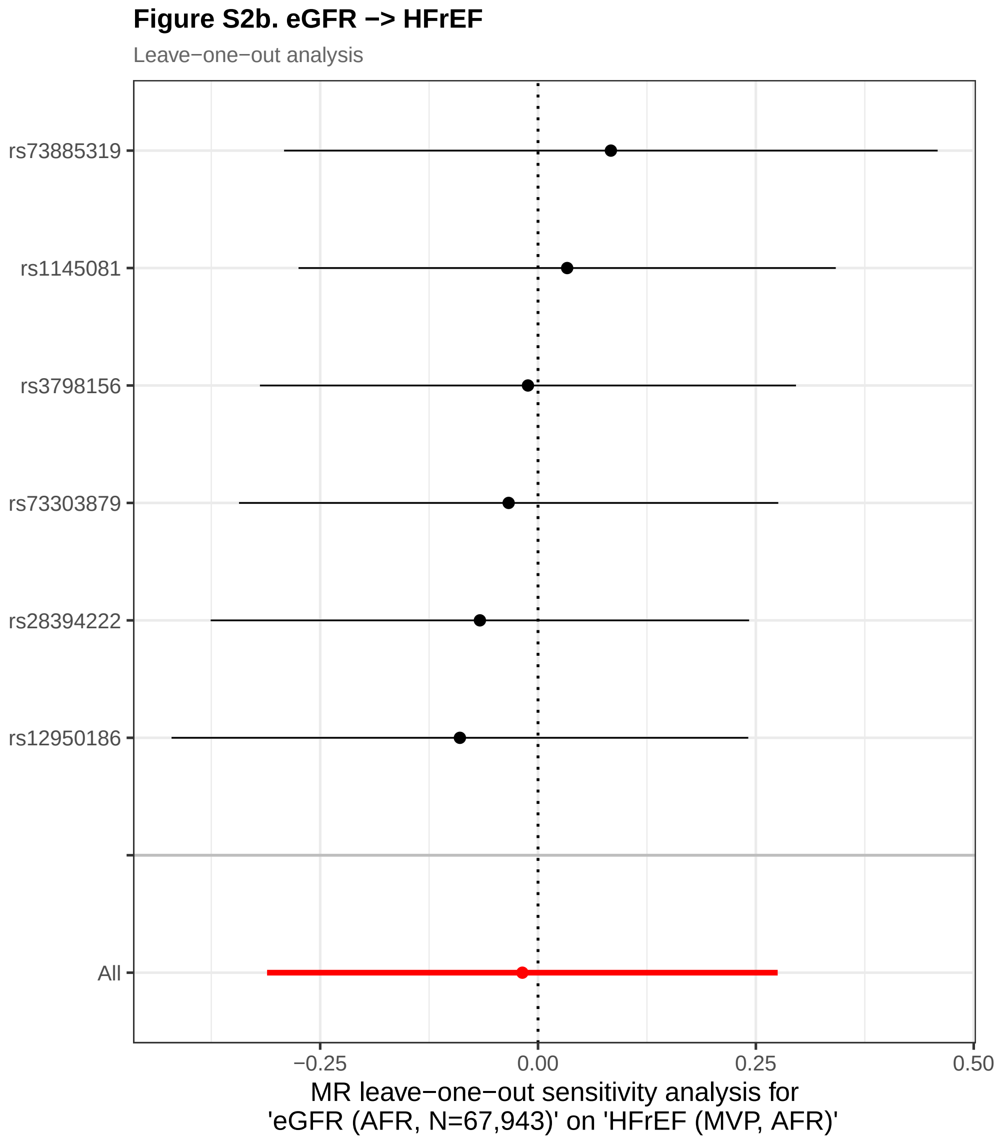


Each row shows the IVW estimate after sequentially removing one SNP instrument.
The bottom row (All) shows the overall IVW estimate using all instruments.
Stability of estimates across rows indicates no single SNP drives the result.
HFpEF, heart failure with preserved ejection fraction; HFrEF, heart failure with reduced ejection fraction.

### Figure S3. Funnel plots of single−SNP MR estimates

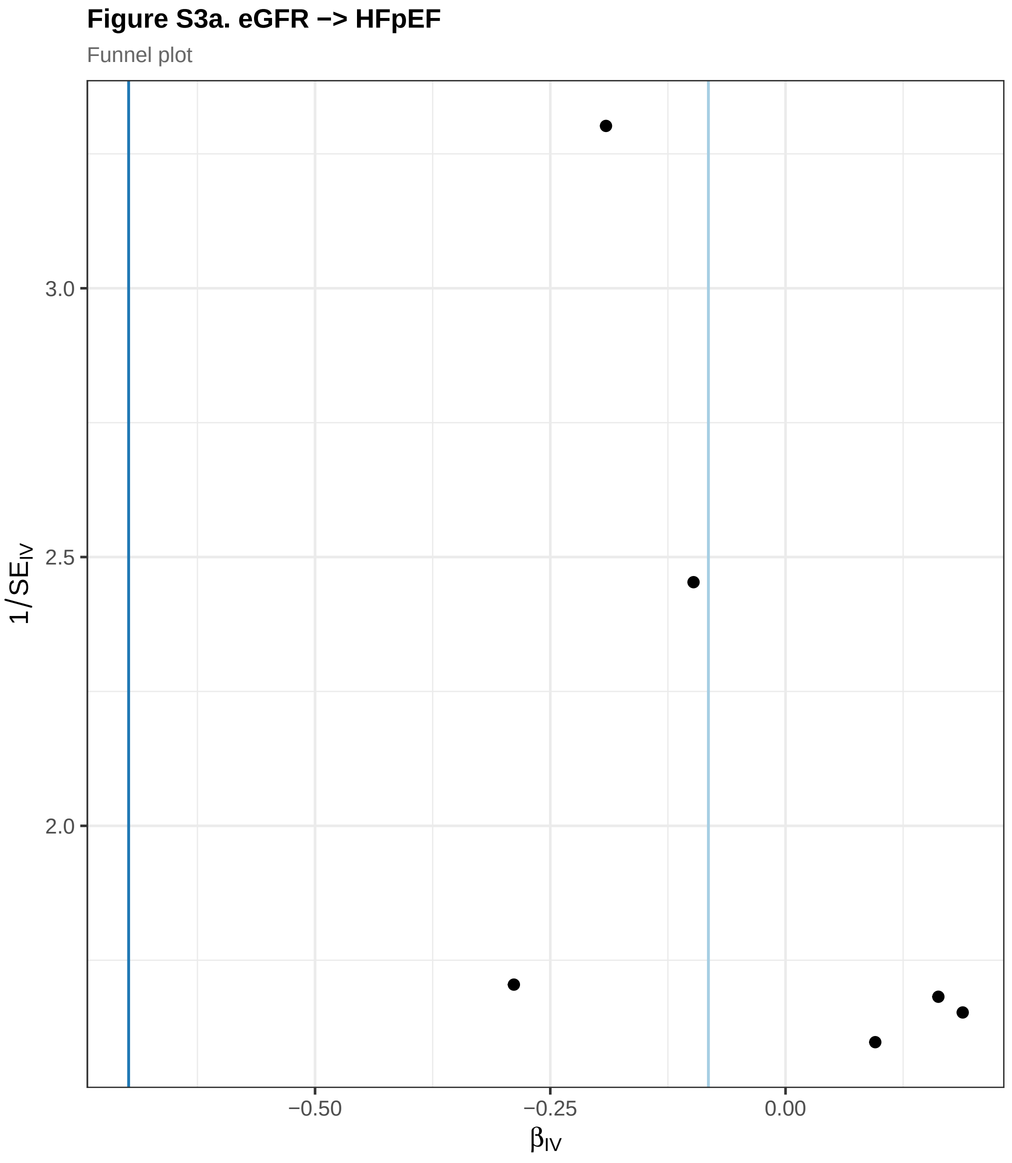


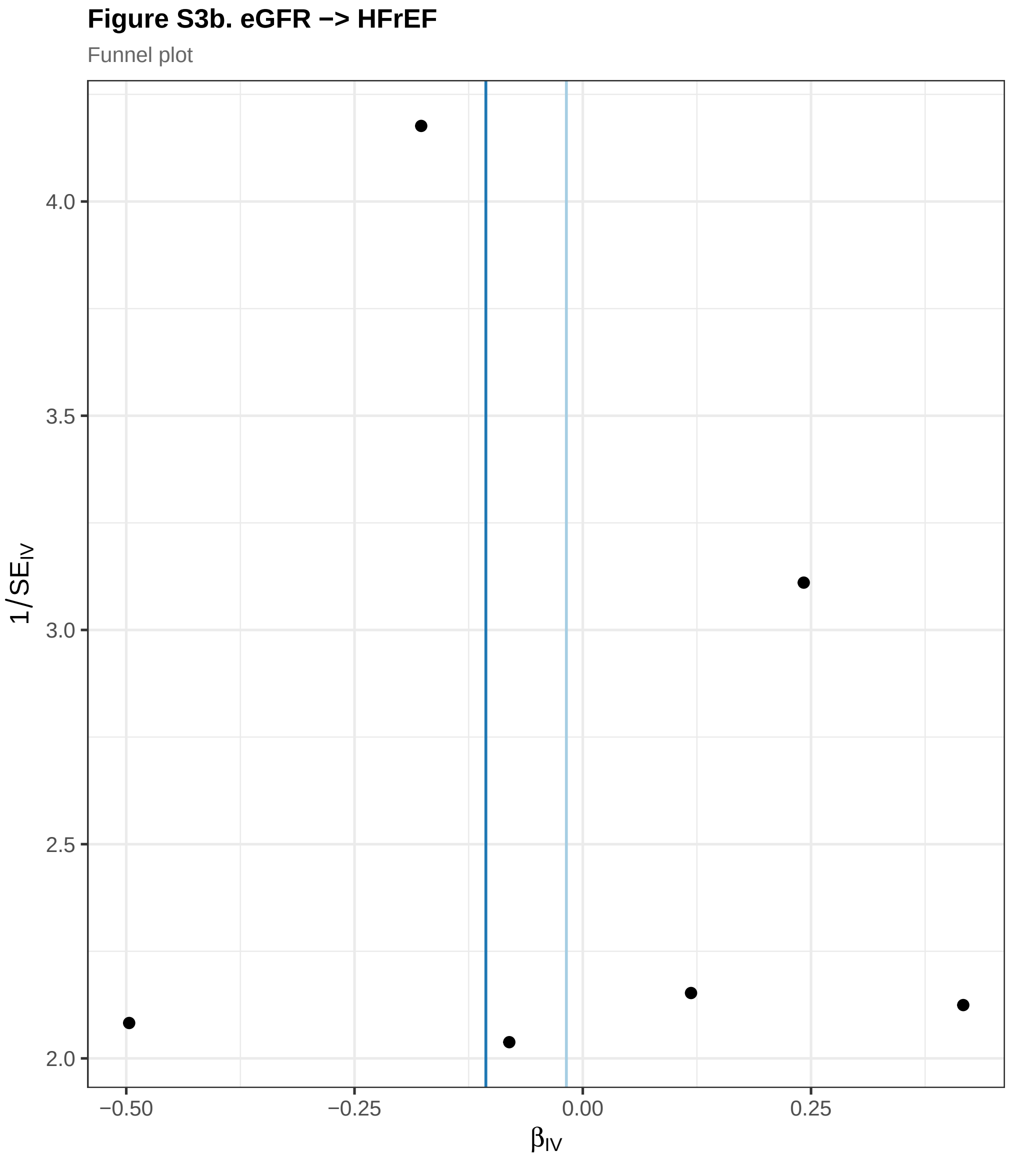


Each point represents one SNP instrument. Vertical lines show IVW (light blue) and MR−Egger (dark blue) pooled estimates.
Symmetry around the pooled estimate suggests absence of directional horizontal pleiotropy.
HFpEF, heart failure with preserved ejection fraction; HFrEF, heart failure with reduced ejection fraction.
