## Supplemental Tables for "An ancestry-matched Mendelian randomisation analysis of kidney function and heart failure subtypes in African ancestry populations"

### Additional file 1: Supplementary Tables

#### Table S1. Steiger filtering results for eGFR genetic instruments

| SNP | Steiger direction | P-value | Retained |
| --- | --- | --- | --- |
| rs1145081 | TRUE | 0.006 | Yes |
| rs12950186 | TRUE | <0.001 | Yes |
| rs28394222 | TRUE | 0.006 | Yes |
| rs3798156 | TRUE | 0.006 | Yes |
| rs73303879 | TRUE | 0.006 | Yes |
| rs73885319 | TRUE | <0.001 | Yes |

*Steiger filtering was applied to verify that each instrument explains greater variance in the exposure (eGFRcrea) than in the outcome, consistent with the hypothesised causal direction. Results were identical for HFpEF and HFrEF outcomes and are presented once. Steiger direction = TRUE indicates the SNP explains more variance in eGFRcrea than in the heart failure outcome. P-value refers to the test of directionality. Retained = Yes indicates the SNP passed filtering and was included in the primary analysis. SNP, single nucleotide polymorphism; eGFRcrea, estimated glomerular filtration rate based on serum creatinine*

#### Table S2. Full MR results — eGFR → HFpEF and HFrEF

Primary method: IVW random-effects (bold). OR, odds ratio per SD increase in genetically predicted eGFRcrea; CI, confidence interval; IVW, inverse-variance weighted. Heterogeneity Q p-value reported for IVW only; MR-Egger intercept p-value reported for MR-Egger only. HFpEF, heart failure with preserved ejection fraction; HFrEF, heart failure with reduced ejection fraction.

| **Outcome** | **Method** | **SNPs (n)** | **OR** | **95% CI** | **P-value** | **Heterogeneity Q p** | **Egger intercept p** |
| --- | --- | --- | --- | --- | --- | --- | --- |
| **HFpEF** | **IVW (random effects)** | **6** | **0.92** | **0.80–1.06** | **0.248** | **0.983** |  |
|  | Weighted median | 6 | 0.88 | 0.56–1.37 | 0.570 |  |  |
|  | MR-Egger | 6 | 0.50 | 0.09–2.61 | 0.455 |  | 0.496 |
|  | Weighted mode | 6 | 0.85 | 0.36–2.00 | 0.720 |  |  |
| **HFrEF** | **IVW (random effects)** | **6** | **0.98** | **0.78–1.23** | **0.878** | **0.693** |  |
|  | Weighted median | 6 | 0.91 | 0.63–1.31 | 0.616 |  |  |
|  | MR-Egger | 6 | 0.90 | 0.24–3.34 | 0.882 |  | 0.899 |
|  | Weighted mode | 6 | 0.90 | 0.41–1.99 | 0.802 |  |  |

#### Table S3. Heterogeneity and pleiotropy diagnostics

Cochran Q statistic from IVW model; MR-Egger intercept test for directional horizontal pleiotropy. SE, standard error of Egger intercept.

| **Outcome** | **Cochran Q** | **df** | **Q p-value** | **Egger intercept** | **SE** | **Intercept p-value** |
| --- | --- | --- | --- | --- | --- | --- |
| HFpEF | 0.704 | 5 | 0.983 | 0.038 | 0.051 | 0.496 |
| HFrEF | 3.045 | 5 | 0.693 | 0.005 | 0.040 | 0.899 |

#### Table S4. Statistical power and minimum detectable effects

MDE calculated at 80% power and α = 0.05. R^2^, total variance in eGFRcrea explained by retained instruments; N_eff_ = 4 / (1/N_cases_ + 1/N_controls_); MDE, minimum detectable effect.

| **Outcome** | **R² (total)** | **Neff** | **MDE (log-OR)** | **MDE (OR)** |
| --- | --- | --- | --- | --- |
| HFpEF (MVP, AFR) | 0.0062 | 20,539 | 0.25 | 1.28 |
| HFrEF (MVP, AFR) | 0.0062 | 33,624 | 0.19 | 1.22 |
